# Under the Microscope: Formaldehyde Exposure in National Health Service Pathology Departments in the United Kingdom

**DOI:** 10.1101/2025.08.22.25333970

**Authors:** Magdalena Plesa, Richard L. Yates

## Abstract

**Objectives:** The United States Environmental Protection Agency has determined that formaldehyde presents an “unreasonable risk of injury to human health.” Occupational inhalation exposure is associated with short- and long-term damage to the respiratory, female reproductive, and nervous systems, and is also carcinogenic. The European Union (EU) has recently introduced formaldehyde workplace exposure limits (WELs) that are lower (long-term: 0.3ppm; short-term: 0.6ppm) than those currently applied in the United Kingdom (UK) (long-term: 2ppm; short-term: 2ppm). UK regulation additionally requires exposure to carcinogens to be reduced to *as low as is reasonably practicable*. We evaluated formaldehyde airborne concentrations in National Health Service (NHS) cell pathology departments to assess the adequacy of exposure controls.

**Methods:** Using the Freedom of Information Act (2000), we requested 12 months (2024–2025) of formaldehyde airborne monitoring data collected by cell pathology departments across n=122 NHS Trusts in England (n=102), Scotland (n=10), Wales (n=6), and Northern Ireland (n=4). Results were evaluated empirically and using EN 689:2018 statistical methods to assess exposure variability, estimate upper-bound concentrations, and determine the likelihood of adequate exposure control when benchmarked against EU WELs.

**Results:** A total of 1,715,516 formaldehyde airborne monitoring results were disclosed by n=117 cell pathology departments. Monitoring was infrequent, with 73% of sites measuring formaldehyde airborne concentrations once weekly or less. EU long-term WELs were exceeded regularly at 70% of sites (95th percentile >0.3 ppm), and EU short-term WELs were exceeded regularly at 43% of sites (95th percentile >0.6 ppm). The 95th percentile upper tolerance limit (UTL_95,70_) exceeded the EU short-term WEL at 68% of sites. Only 11% and 17% of departments demonstrated frequent (once daily or more) formaldehyde airborne monitoring with 95th percentiles below the EU long- and short-term WELs, respectively.

**Conclusions:** Formaldehyde exposure is infrequently monitored and inadequately controlled in NHS cell pathology departments.

**What is already known:** A substantial body of occupational exposure data shows that formaldehyde inhalation is associated with myriad short- and long-term deleterious health effects on the respiratory, female reproductive, and nervous systems. It is also a human carcinogen. Pathology departments are amongst the riskiest occupational environments for formaldehyde inhalation exposure and therefore require a high standard of governance and infrastructure to adequately protect staff.

**What this study adds:** We show that formaldehyde airborne concentrations in most NHS cell pathology departments are monitored infrequently and regularly exceed EU WELs. Our data raises concern for the health of thousands of NHS employees working in these environments.

**How this study might affect research, practice, or policy:** Urgent national regulatory intervention is now warranted to improve the occupational hygiene of NHS cell pathology departments. This will require a combination of upgraded infrastructure, more regular personal exposure monitoring, better employee education on basic lab practice and occupational health risks, improved access to appropriate personal protective equipment, management accountability for occupational health, and external oversight by the Health and Safety Executive.

## Introduction

The United States Environmental Protection Agency (EPA) completed its updated risk evaluation for formaldehyde in 2024 and determined that it presents an “unreasonable risk of injury to human health”.^1^ Occupational formaldehyde inhalation causes mild to severe eye and respiratory tract irritation,^2^ decreases pulmonary function,^3,4^ induces respiratory tract histopathological lesions,^5–7^ and increases the prevalence and severity of allergic conditions and asthma.^8,9^ Beyond the respiratory system occupational formaldehyde inhalation has been associated with an increased time-to-pregnancy and risk of spontaneous abortion,^10^ the motor neuron disease amyotrophic lateral sclerosis,^11–13^ and global cognitive impairment.^14^ Chronic formaldehyde inhalation is also carcinogenic to humans according to the International Agency for Research on Cancer.^15^ Sufficient evidence now supports a causal relationship^16^ with nasopharyngeal carcinoma,^17^ sinonasal carcinoma,^18^ and myeloid leukaemia.^19^ Hypopharyngeal carcinoma,^20^ Hodgkin lymphoma,^21^ and multiple myeloma^21^ are also associated.

Due to increasing concern of formaldehyde’s toxicity, the European Union (EU) Scientific Committee on Occupational Exposure Limits recommended in 2016 that formaldehyde work exposure limits (WELs) be revised to 0.3ppm (8 hour time-weighted average) and 0.6ppm (15 minute short-term exposure limit).^22^ These recommendations were ratified in a 2019 EU mandate^23^ that had a mandatory transposition date into EU member state law of July 2021.^23^ A transition period for the healthcare sector ended in July 2024.^23^ The UK left the EU in 2020 and was not obligated to adopt this updated regulation. The current UK formaldehyde WELs^24^ are the world’s highest defined permissible occupational limits (UK short-term WEL: 2ppm; UK long-term WEL: 2ppm) (Fig. 1).

**Figure 1:**
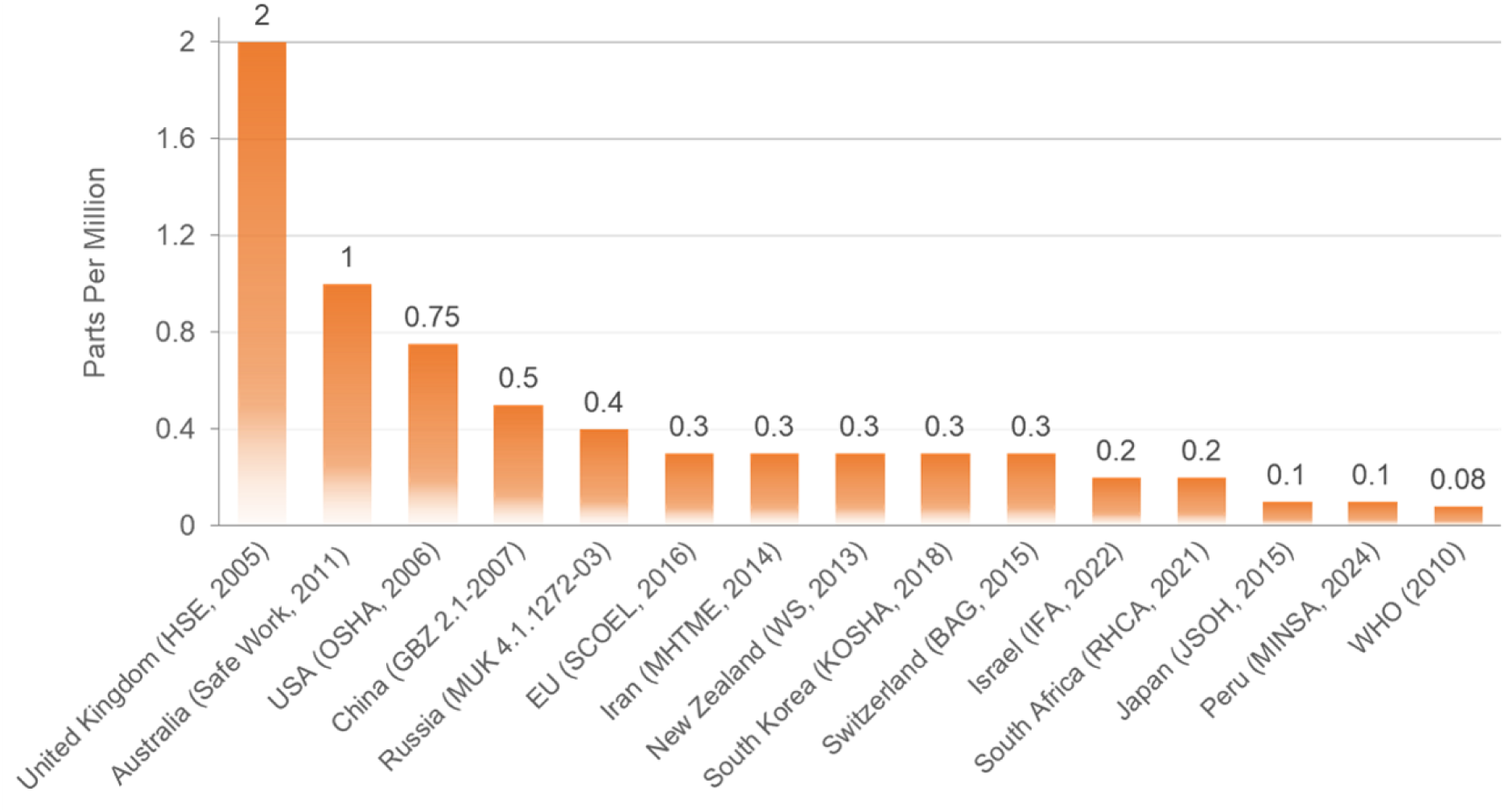
Comparison of International Formaldehyde Work Exposure Limits (8 hour time-weighted average). OSHA = Occupational Safety and Health Administration. EU = European Union. SCOEL = Scientific Committee on Occupational Exposure Limits. MHTME = Ministry of Health, Treatment and Medical Education. WS = Worksafe. KOSHA = Korea Occupational Safety and Health Agency. BAG = Bundesamt für Gesundheit. IFA = Institut für Arbeitsschutz. RHCA = Regulations for Hazardous Chemical Agents. JSOH = Japan Society for Occupational Health. MINSU = Ministerio de Salud. WHO (World Health Organisation) recommendation represents an indoor air limit and is added for comparison.

Formaldehyde is designated a Class 1B carcinogen under UK classification and labelling regulations.^25^ The Control of Substances Hazardous to Health Regulations 2002 (COSHH) state that control of formaldehyde exposure as a carcinogen is defined as adequate only when (a) the principles of good control practice are applied, (b) UK WELs are not exceeded, *and* (c) exposure is reduced to *as low as is reasonably practicable* (ALARP).^26^ The principle of ALARP mandates that formaldehyde exposure be reduced to such an extent that any further reduction would require disproportionate sacrifice when weighed against the health risks (which should be insignificant).^27^ A substantial body of evidence now associates occupational formaldehyde inhalation with health risks even at relatively low exposures that are significantly below the UK WEL.^28^ Revised EU regulation therefore provides an empirical benchmark for the evaluation of formaldehyde airborne concentrations in UK workplaces that may be considered *reasonably practicable*.

Pathology services in the UK employ over 28,000 people in England alone.^29^ Pathology laboratories are amongst the riskiest occupational settings for inhalation exposure to formaldehyde where it is used in a buffered solution (formalin) to preserve human tissue.^30^ Deleterious health effects in pathology laboratory workers are already well described,^31^ and include DNA damage that may be associated with cancer morbidity.^32–37^ The extent of formaldehyde exposure amongst UK pathology laboratory workers employed by the National Health Service (NHS) has not been investigated previously. The UK Freedom of Information Act (2000) allows access to environmental monitoring data where it is held by a public body.^38^ This provides a unique opportunity to investigate formaldehyde airborne concentrations in NHS cell pathology departments. Our data provides evidence that formaldehyde is infrequently monitored and poorly controlled in the NHS, raising serious concern for the health of thousands of employees that now warrants urgent national regulatory intervention.

## Methods

### Freedom of Information (2000) Act Request

Under the UK Freedom of Information Act (2000), we requested 12 months (2024-25) of formaldehyde airborne monitoring results collected as part of the regular scheduled area monitoring protocols of cell pathology departments in n=122 NHS Trusts in England (n=102), Scotland (n=10), Wales (n=6), and Northern Ireland (n=4). Every NHS Trust that documented a cell pathology department on their website was included in the request.

We asked NHS Trusts to disclose on how many occasions in the preceding 12 months their cell pathology department(s) had recorded formaldehyde airborne concentrations that were (a) >0ppm and ≤0.3ppm, (b) >0.3ppm and ≤0.4ppm, (c) >0.4ppm and ≤0.5ppm, (d) >0.5ppm and ≤0.6ppm, (e) >0.6ppm and ≤0.7ppm, (f) >0.7ppm and ≤0.8ppm, (g) >0.8ppm and ≤0.9ppm, (h) >0.9ppm and ≤1ppm, (i) >1ppm and ≤1.5ppm, (j)>1.5ppm and ≤2ppm and (k) >2ppm. Bin (k) was required to be open-ended to ensure all formaldehyde air monitoring data held by an NHS Trust was disclosed.

We also asked how often scheduled formaldehyde airborne monitoring was undertaken, the total number of times formaldehyde airborne monitoring had been undertaken in the preceding 12 months, and the approximate annual caseload of surgical specimens.

The Freedom of Information Act (2000) contains provisions that allow public bodies to refuse access to records if it is felt gathering, reviewing or preparing requested documents would take an excessive amount of time and effort.^38^ The Freedom of Information request used in the current study was therefore intended to balance the level of detail required for a worthwhile investigation whilst at the same time avoiding a significant number of NHS Trusts applying an exemption and refusing to disclose data. Further, due to many NHS Trusts relying on paper records to document formaldehyde air monitoring, we did not request raw data as this would have led to many refusals owing to the time needed to scan potentially thousands of documents.

### Frequency of Scheduled Formaldehyde Airborne Monitoring

The frequency of scheduled formaldehyde airborne monitoring was defined as follows: Continuous (at least once-hourly); Daily (at least once-daily); Weekly (at least once-weekly); Monthly (at least once 3-weekly); Quarterly (at least once 3-monthly); Annually (at least once 9-monthly).

### Empirical Analysis of Formaldehyde Airborne Concentrations and Comparison to EU and UK WELs

The proportion (%) of formaldehyde airborne monitoring data exceeding EU and UK WELs was calculated for each cell pathology department and averaged across the cohort. Data is presented as the mean ± SEM. Percentile estimates of formaldehyde airborne concentrations were calculated using Monte Carlo simulation with a uniform distribution to avoid potential bias from assuming parametric forms. The analysis was performed using Python 3.12 with NumPy and Pandas libraries. The Monte Carlo algorithm proceeded as follows: for each iteration, random samples were drawn uniformly from within each bin according to the observed frequencies, creating a reconstructed dataset. The 50^th^ and 95^th^ percentiles of formaldehyde airborne concentrations were then calculated from this reconstructed dataset. This process was repeated for 1,000 iterations to generate a distribution of percentile estimates. Point estimates were calculated as the mean of the Monte Carlo iterations, and 95% confidence intervals were determined for each percentile estimate using the 2.5th and 97.5th percentiles of the Monte Carlo distribution.^39^

To confirm the validity of our findings, we performed a sub-group analysis comparing the proportion (%) of airborne formaldehyde monitoring data exceeding EU and UK WELs between sites with frequent monitoring (continuous or daily) and sites with infrequent monitoring (weekly, monthly, quarterly, or annually). This sub-group analysis was also replicated with the derived percentile data.

Sites where both the empirical 50^th^ and 95^th^ percentiles of formaldehyde airborne concentration were estimated to be below 0.3ppm were not assessed further. This avoided artefactual over precision when variability of frequency data was limited at sites with the lowest airborne concentrations and ensured a conservative methodology was maintained that was weighted towards compliance.

### Evaluation of Airborne Formaldehyde Concentrations using EN 689:2018

Cell pathology departments in which the empirical 95^th^ percentile of formaldehyde airborne concentration was >0.3ppm were further evaluated in line with the statistical approach described in Annex F (*Statistical test for testing compliance with occupational exposure limit value)* of EN 689:2018 (*Measurement of exposure by inhalation to chemical agents - Strategy for testing compliance with occupational exposure limit values*). EN 689:2018 is the current EU and UK standard for measuring workplace inhalation exposure to chemical agents.^40^

As described in EN689:2018, the geometric mean (GM) and geometric standard deviation (GSD) of formaldehyde airborne concentration for each cell pathology department were calculated using log-transformed data. The outcome metric (UTL_95,70_) was then calculated using the formula:

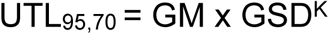

The tolerance factor *k* represents the normal deviate for the 95^th^ percentile added to an uncertainty factor related to sample size that was derived from published tabulated values in BS689:2018 (Annex F, Table F.1).

The UTL_95,70_ represents a statistical estimate of the 95th percentile of airborne formaldehyde concentrations. It is calculated such that there is 70% confidence that the true 95th percentile does not exceed the UTL_95,70_. It provides a conservative upper-bound estimate of potential concentrations that accounts for both variability in measurement and the uncertainty associated with a finite sample size. The UTL_97,70_ was compared to the EU short-term WEL (0.6ppm) to provide an empirical benchmark of adequate control.

### Combined Assessment of Monitoring Frequency and Airborne Formaldehyde Concentration

Adequate monitoring and control of formaldehyde exposure was considered where there was (A) at least once-daily formaldehyde airborne monitoring showing (B) a 95^th^ percentile <0.3ppm. The proportion of sites with (A) at least once-daily formaldehyde airborne monitoring showing (B) a 95^th^ percentile <0.6ppm was also calculated.

## Results

### Study population

Overall response rate was 100% (122/122). In total, 85% (104/122) of NHS Trusts were able to disclose a suitable 12-month record (2024-25) of formaldehyde airborne monitoring on behalf of n=117 cell pathology departments in England (n=100), Scotland (n=9), Wales (n=5), and Northern Ireland (n=3). In contrast, 15% (18/122) of NHS Trusts could not disclose a suitable record of formaldehyde monitoring (England: n=15; Scotland: n=1; Wales: n=1; Northern Ireland: n=1). Of the n=117 cell pathology departments able to disclose suitable data, 6% (7/117) were specialist hospitals encompassing children’s, women’s, orthopaedic, and ophthalmologic services. The remainder (94%; 110/117) were non-specialist. Across the whole cohort, 1,715,516 distinct monitoring events were disclosed.

The average annual caseload of surgical specimens reported by departments with a suitable record of formaldehyde monitoring was 36,959 (range: 1,780 – 99,627; standard deviation = 22,777).

NHS Trusts could not disclose suitable data for the following reasons: n=5 NHS Trusts applied Section 12 of the Freedom of Information Act (cost of compliance exceeds the appropriate limit); n=3 NHS Trusts stated they did not have records available to the level of detail requested; n=6 NHS Trusts disclosed incomplete or missing data that precluded analysis; n=2 NHS Trusts only documented formaldehyde monitoring when 2ppm was exceeded which precluded analysis; and n=2 NHS Trusts refused to disclose data as their laboratory was managed by a third party.

### Frequency of Scheduled Formaldehyde Airborne Monitoring

Of the NHS cell pathology departments that were able to disclose a suitable record formaldehyde airborne monitoring, 16% (19/117) monitored formaldehyde airborne concentrations continuously, 11% (13/117) monitored formaldehyde airborne concentrations daily, 19% (22/117) monitored formaldehyde airborne concentrations weekly, 34% (40/117) monitored formaldehyde airborne concentrations monthly, 15% (18/117) monitored formaldehyde airborne concentrations quarterly, and 4% (5/117) monitored formaldehyde airborne concentrations annually (Fig. 2).

**Figure 2:**
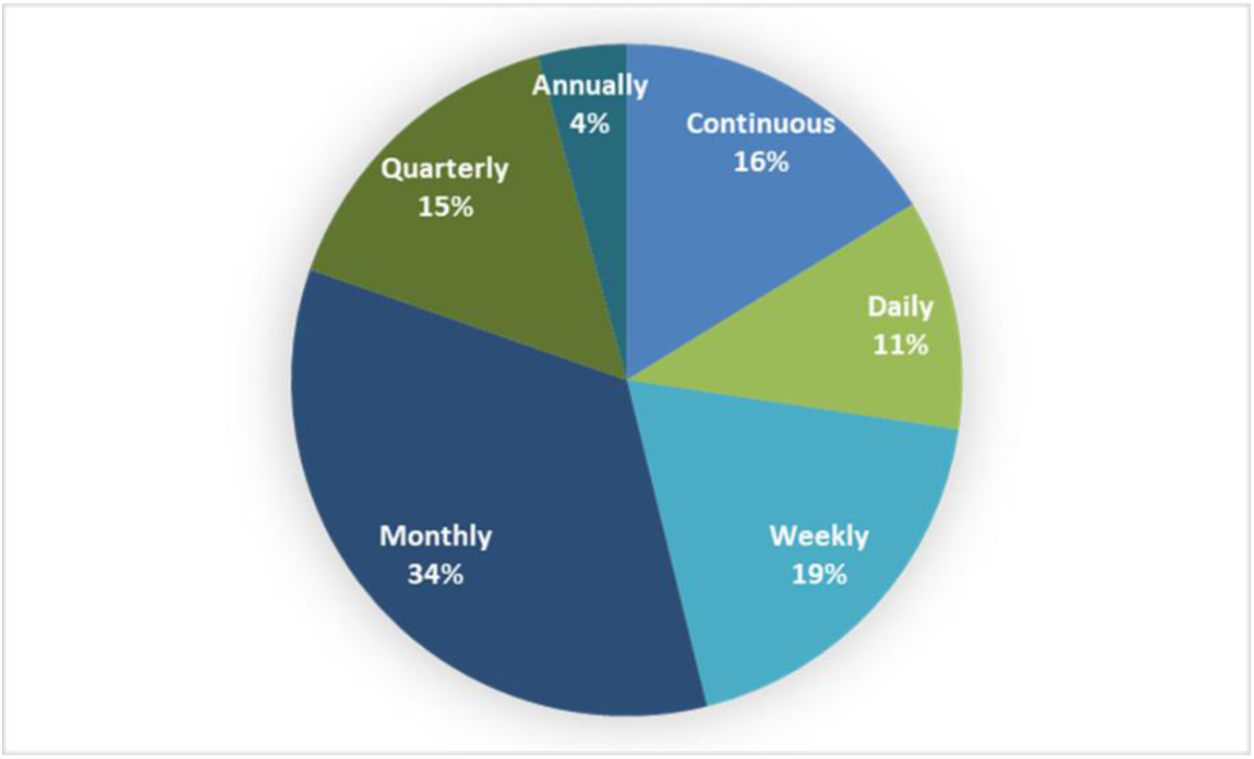
Frequency of Scheduled Formaldehyde Airborne Monitoring in NHS Cell Pathology Departments.

The number of distinct monitoring events disclosed by NHS Trusts varied widely between sites and was related to both the frequency of regular airborne monitoring and the number of areas within each department assessed during monitoring schedules. For departments with continuous monitoring the average number of disclosures was 89,008 ± 33,640 (mean ± SEM). For departments with daily monitoring the average number of disclosures was 936 ± 255 (mean ± SEM). For departments with weekly monitoring the average number of disclosures was 364 ± 182 (mean ± SEM). For departments with monthly monitoring the average number of disclosures was 90 ± 22 (mean ± SEM). For departments with quarterly monitoring the average number of disclosures was 29 ± 7 (mean ± SEM). For departments with annual monitoring the average number of disclosures was 13 ± 6 (mean ± SEM).

### Empirical Analysis of Formaldehyde Airborne Concentrations and Comparison to EU and UK WELs

Across the whole cohort (n=117), the average proportion of annual formaldehyde airborne monitoring at each site that exceeded the EU long-term WEL (0.3ppm) was 23.6% ± 2.3%. The average proportion of formaldehyde airborne monitoring at each site that exceeded the EU short-term WEL (0.6ppm) was 8.5% ± 1.2%. The average proportion of formaldehyde airborne monitoring at each site that exceeded the UK short- and long-term WEL (2ppm) was 0.3% ± 0.1%.

Empirical percentile analyses showed that 70% (82/117) of NHS cell pathology departments had a 95^th^ percentile of formaldehyde airborne concentrations exceeding the EU long-term WEL (0.3ppm) and 43% (50/117) of NHS cell pathology departments had a 95^th^ percentile of formaldehyde airborne concentrations exceeding the EU short-term WEL (0.6ppm). Nil (0/117) NHS cell pathology departments had a 95^th^ percentile of formaldehyde airborne concentrations that exceeded the UK short- and long-term WEL (2ppm). However, almost a third of sites (30%; 35/117) had recorded an airborne formaldehyde concentration exceeding 2ppm at least once in the preceding 12 months. Empirical percentile analyses of formaldehyde airborne concentrations are presented in Fig. 3.

**Figure 3:**
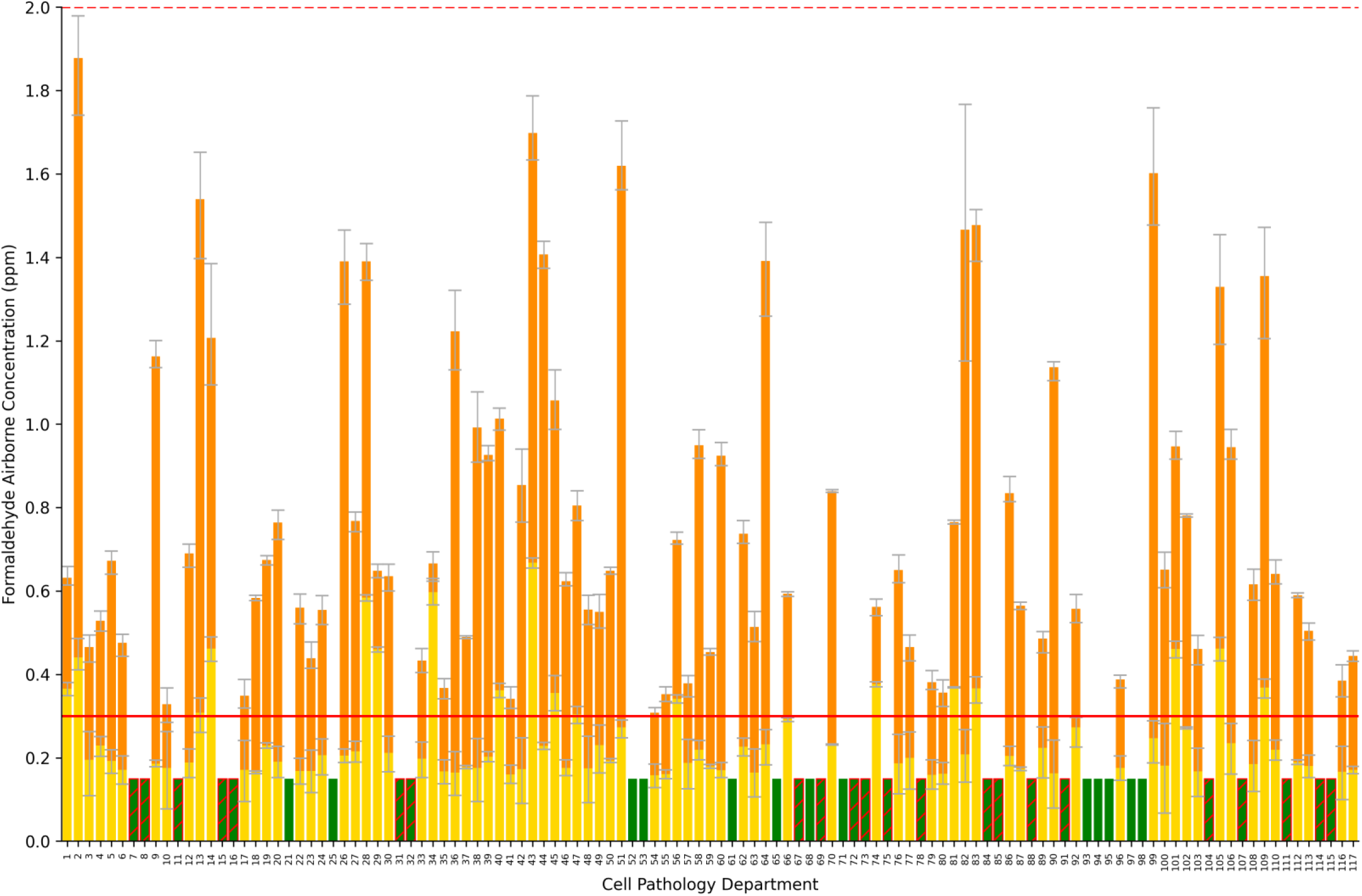
Comparison of Formaldehyde Airborne Concentrations in NHS Cell Pathology Departments with EU and UK Long-Term WELs. The graph depicts the 50th percentile (yellow bars) and the distance from the 50th percentile to the 95th percentile (orange bars) of formaldehyde airborne concentrations for each NHS cell pathology department that disclosed a suitable record of monitoring. Error bars represent the 95% confidence intervals of each percentile estimate. The overlaid red horizontal line depicts the EU long-term WEL (0.3ppm). The overlaid broken red line depicts the UK long-term WEL (2ppm). Green bars represent cell pathology departments where both the 50th and 95th percentiles of formaldehyde airborne concentrations were <0.3ppm. Precise percentile estimates below 0.3ppm could not be generated. Green bars with red strikethroughs represent departments where both the 50th and 95th percentiles of formaldehyde exposure monitoring were <0.3ppm but that reported infrequent monitoring (once weekly or less).

In our sub-group analysis, results did not differ significantly between frequent monitoring (continuous or daily; n=32) and infrequent monitoring (weekly, monthly, quarterly, and annually; n=85) with respect to the average proportion of airborne monitoring >0.3ppm (infrequent: 23.9 ± 2.6% vs. frequent: 22.9 ± 5.0%, p=0.849), >0.6ppm (infrequent: 8.6 ± 1.4% vs. frequent: 8.3 ± 2.5%, p=0.892), or >2ppm (infrequent: 0.3 ± 0.1% vs. frequent: 0.2 ± 0.07%, p=0.428). Results also did not differ significantly between infrequent and frequent monitoring with respect to the proportion of sites with a 95^th^ percentile of airborne monitoring >0.3ppm (infrequent: 74% vs. frequent: 59%, p=0.123), 95^th^ percentile of airborne monitoring >0.6ppm (infrequent: 45% vs. frequent: 38%, p=0.487), 95^th^ percentile of airborne monitoring >2ppm (infrequent: 0% vs. frequent: 0%, p=1.0). There was a significant difference in the proportion of sites that reported formaldehyde airborne monitoring >2ppm at least once in the last 12 months (infrequent monitoring: 19% vs. frequent monitoring: 59%, p=<0.001).

### Evaluation of Airborne Formaldehyde Concentrations using EN 689:2018

In total, n=81/117 cell pathology departments met criteria to be submitted for evaluation using EN 689:2018. The upper tolerance limit of the 95^th^ percentile (UTL_95,70_) exceeded the EU short-term WEL (0.6ppm) at 97.5% of assessed sites (79/81). These sites represent 68% of the whole cohort (79/117).

### Combined Assessment of Monitoring Frequency and Airborne Formaldehyde Concentration Distribution

Of the NHS cell pathology departments that disclosed a suitable record of formaldehyde airborne monitoring data, only 11% (13/117) had daily or continuous monitoring demonstrating a 95th percentile lower than the EU long-term WEL (0.3ppm) and only 17% (20/117) had daily or continuous monitoring demonstrating a 95th percentile lower than the EU short-term WEL (0.6ppm).

## Discussion

We present a comprehensive assessment of formaldehyde airborne concentrations in NHS cell pathology departments throughout the UK using data now publicly available and verifiable. Over 1.7 million distinct formaldehyde airborne monitoring results have been disclosed from 117 NHS cell pathology departments in England (n=100), Scotland (n=9), Wales (n=5), and Northern Ireland (n=3) covering a period of 12 months from 2024 to 2025. Our data shows that most sites monitor formaldehyde airborne concentrations infrequently (once weekly or less at 73% of sites), despite handling tens of thousands of surgical specimens annually, and that formaldehyde exposure is poorly controlled. Many cell pathology departments report airborne formaldehyde concentrations that regularly exceed the EU long- and short-term WELs (95th percentile > 0.3ppm at 70% of sites; 95th percentile > 0.6ppm at 43% of sites). Evaluation of formaldehyde airborne monitoring results using EN 689:2018 provides further robust statistical evidence suggestive of poor control (UTL_95,70_ > EU short-term WEL at 68% of sites). Our data warrants urgent national regulatory intervention to improve the infrastructure and governance of NHS cell pathology departments and protect the health of thousands of NHS employees.

Formaldehyde is a sensory irritant that induces mild to severe symptoms upon acute airborne exposure including stinging, burning, itching, watering eyes, rhinitis, sneezing, coughing, sore throat, and bronchial constriction.^28^ Controlled human exposure studies have shown that irritant symptoms begin at formaldehyde inhalation of approximately 0.3ppm and increase in severity with airborne concentration.^2^ Occupational exposure studies demonstrate that formaldehyde inhalation causes decrements in pulmonary function,^3,4^ induces histopathological lesions of the respiratory tract,^5–7^ and increases the incidence and severity of allergy and asthma.^8,9^ The US EPA have recently defined the lifetime exposure reference concentration for formaldehyde’s respiratory system non-cancer effects as 0.007mg/m^3^, 357 times lower than the current UK WEL.^28^ The toxicity of formaldehyde is not limited to the respiratory tract. Occupational exposure studies have also shown that formaldehyde inhalation is associated with an increased time-to-pregnancy and risk of spontaneous abortion,^10^ with the relevant period of formaldehyde exposure potentially extending well before conception.^28^ The US EPA lifetime exposure reference concentration for formaldehyde’s female reproductive effects is 0.01mg/m^3^, 247 times lower than the current UK WEL.^28^ Emerging evidence from multiple occupational cohorts has also associated formaldehyde inhalation exposure with the motor neuron disease amyotrophic lateral sclerosis,^11–13^ as well as global cognitive impairment.^14^

Formaldehyde is designated a Group 1 carcinogen by the International Agency for Research on Cancer.^15^ Multiple human cohort and case-control studies now provide sufficient evidence^16^ that chronic formaldehyde inhalation causes nasopharyngeal^17^ and sinonasal carcinoma,^18^ and myeloid leukaemia.^19^ Hypopharyngeal carcinoma,^20^ Hodgkin lymphoma,^21^ and multiple myeloma^21^ have also been associated. Formaldehyde has been shown to be a direct-acting genotoxic agent in a variety of *in vitro* systems^41^, in experimental animal models (mice,^42^ rats,^43^ and monkeys^44^) and in humans.^32–37^ Specifically, formaldehyde has been shown to induce point mutations,^45^ deletions,^45^ DNA-DNA^46^ and DNA-protein^41^ cross-links,^47^ DNA mono adducts,^47^ chromosomal aberration^35^ and micronuclei formation,^34^ single-strand^48^ and double-strand breaks,^49^ sister chromatid exchanges,^50^ inhibition of unscheduled DNA synthesis,^48^ DNA repair inhibition,^48^ and cellular transformation.^51^ Aneuploidy in chromosomes 1,5, and 7 in circulating myeloid progenitor cells, a primary target for lymphohematopoietic carcinogenesis, has also been reported^52^ as well as disrupted haematopoiesis.^53^ Multiple studies have demonstrated genotoxic damage in both buccal cells and peripheral blood leukocytes of pathology lab workers at relatively low exposure concentrations.^32–37^ A very high standard of infrastructure and governance is therefore required to adequately protect employees working in these environments.

Under UK COSHH regulation, formaldehyde as a carcinogen is defined as adequately controlled only if (a) the principles of good control practice are applied, (b) UK WELs are not exceeded, *and* (c) exposure is reduced to *as low as is reasonably practicable* (ALARP).^26^ ALARP mandates that exposure to carcinogens be reduced such that any further reduction would require disproportionate sacrifice when weighed against the health risks (which should be insignificant).^27^ A substantial body of evidence demonstrates that formaldehyde is associated with myriad deleterious health effects at concentrations well below UK WELs.^28^ Revised EU WELs therefore set an important new precedent for UK employers. It is difficult to argue that the time, effort, and cost required to reduce formaldehyde exposure to below 0.3ppm requires disproportionate sacrifice in the UK when this benchmark is already mandated in 27 EU member states. We believe, therefore, that there are strong scientific and regulatory arguments for mandating at least once-daily monitoring of formaldehyde airborne concentrations showing a 95^th^ percentile lower than the EU long-term WEL (0.3ppm). Our data shows that, currently, 89% of NHS cell pathology departments fail to conform to this standard and 83% fail to conform even when the less stringent EU short-term WEL (0.6ppm) is used. Our findings raise concern that formaldehyde exposure is not adequately controlled in NHS cell pathology departments in line with ALARP and underscore the importance of implementing updated UK WELs consistent with EU regulation (Directive (EU) 2019/983) to better protect employees.^23^

We believe significant advances can be made to improve the occupational hygiene of NHS cell pathology departments without the need for disproportionate investment.^54–56^ Evaluation of formaldehyde exposure in pathology departments elsewhere in the world has shown typically lower concentrations than we report, including recently in Iran,^57,58^ Italy,^59^ and Japan.^56^ Our data shows large differences in the 50^th^ and 95^th^ percentiles of formaldehyde airborne concentrations suggesting that it is high-exposure tasks that are poorly controlled and that should be targeted for intervention. Inhalation exposure to formaldehyde in NHS cell pathology departments is mostly controlled by local exhaust ventilation, with open down-draught tables widely employed on which high-exposure tasks are undertaken. These units provide partial exposure control that is variable, not well-defined, and not apparent to the user in real time. Our data suggests that the current reliance on this infrastructure is not providing a high standard of exposure control in most NHS cell pathology departments. This is likely due to the dilapidated nature of many sites that precludes effective ventilation even when down-draught tables are operating at their full effectiveness.^60^ Purpose built enclosed cabinets with appropriate ventilation and ergonomic capability for safe and effective dissection of large specimens are commercially available and widely used elsewhere.^61^ UK COSHH regulations already mandate the total enclosure of work with formaldehyde where this is reasonably practicable.^62^ Improved personal exposure monitoring, better employee education on basic lab practice and occupational health risks, improved access to appropriate personal protective equipment, management accountability for occupational health, and external oversight by the Health and Safety Executive, are also now warranted.

### Limitations

The data presented is derived from information disclosed by NHS Trusts under the UK Freedom of Information Act (2000) that has not been directly collected by the authors. UK employers are mandated to maintain records of environmental monitoring for at least 5 years. On this basis the data disclosed is reasonably assumed to be robust and reliable. Any discrepancy owing to human error would be expected to be rare and to not overtly affect the findings of the study given the large number of disclosures analysed. It is noted from our findings that monitoring practices vary widely between NHS cell pathology departments and is often infrequent. Unfortunately, we do not have information on the standard operating procedures or equipment used by each disclosing NHS Trust. However, that our data shows consistent elevated airborne formaldehyde concentrations across many cell pathology departments over a 12-month period urges against significant methodological and/or technical differences between sites. We recognise that sampling duration strongly influences the statistical distribution of measured concentrations, with shorter-duration measurements more likely to capture transient peaks, resulting in greater variability and a higher frequency of readings above permissible limits compared with longer averaging times. For this reason, workplace exposure limits have been used solely as empirical benchmarks to aid interpretation of measured concentrations and to provide insight into the potential adequacy of control with respect to the ALARP principle, rather than as indicators of formal compliance with work exposure limits or cumulative exposure of individual employees. We also recognise that intermittent monitoring warrants interpretative caution. However, sporadic area measurements demonstrating elevated exposures are cause for concern given that in indoor environments the half-life of formaldehyde can be measured in hours when there are ongoing sources and poor ventilation.^63^

### Conclusions

The International Labour Organization, of which the UK is a signatory, added “a safe and healthy working environment” to its fundamental principles and rights at work in 2022.^64^ In December 2024, the US EPA released its updated risk evaluation for formaldehyde and determined that it presents an “unreasonable risk of injury to human health”.^1^ The UK has the world’s highest defined formaldehyde WELs.^24^ Nonetheless, the additional regulatory requirement for carcinogenic exposure to be reduced to *as low as is reasonably practicable* should be protecting employees from formaldehyde.^26^ Our data provides evidence that most NHS cell pathology departments monitor formaldehyde infrequently and that staff are regularly exposed to airborne concentrations associated with deleterious health effects. The relevance of our data is, however, not limited to healthcare environments. Industries with occupational exposure to formaldehyde also include manufacturing, construction, and myriad others that employ many tens of thousands of people in the UK. Improved epidemiological studies that examine health outcomes in these environments are now desperately needed alongside urgent regulatory intervention to proactively protect the health of people working with formaldehyde.

## Contributions

MP and RLY contributed equally to the manuscript

## Acknowledgments

The authors wish to thank Dr Christopher Partlett (Assistant Professor of Medical Statistics and Clinical Trials, University of Nottingham, UK) for his assistance with the statistical analyses used in this manuscript.

## Patient Consent for Publication

Not applicable

## Ethics Approval

Not applicable

## Data Availability Statement

Data disclosed under the Freedom of Information Act (2000) is publicly available.

## References

1. US Environmental Protection Agency. Risk Evaluation for Formaldehyde Available from: https://www.epa.gov/assessing-and-managing-chemicals-under-tsca/risk-evaluation-formaldehyde. 2024

2. Lang I, Bruckner T, Triebig G. Formaldehyde and chemosensory irritation in humans: a controlled human exposure study. Regul Toxicol Pharmacol RTP. 2008;50:23–36.

3. Alexandersson R, Hedenstierna G. Pulmonary function in wood workers exposed to formaldehyde: a prospective study. Arch Environ Health. 1989;44:5–11.

4. Uthiravelu P, Saravanan A, Kumar CK, Vaithiyanandane V. Pulmonary function test in formalin exposed and nonexposed subjects: A comparative study. J Pharm Bioallied Sci. 2015;7:S35–S39.

5. Edling C, Hellquist H, Odkvist L. Occupational exposure to formaldehyde and histopathological changes in the nasal mucosa. Br J Ind Med. 1988;45:761–765.

6. Holmström M, Wilhelmsson B, Hellquist H, Rosén G. Histological changes in the nasal mucosa in persons occupationally exposed to formaldehyde alone and in combination with wood dust. Acta Otolaryngol (Stockh). 1989;107:120–129.

7. Boysen M, Zadig E, Digernes V, Abeler V, Reith A. Nasal mucosa in workers exposed to formaldehyde: a pilot study. Br J Ind Med. 1990;47:116–121.

8. Sakamoto T, Doi S, Torii S. Effects of formaldehyde, as an indoor air pollutant, on the airway. Allergol Int. 1999;48:151–160.

9. Lam J, Koustas E, Sutton P, et al. Exposure to formaldehyde and asthma outcomes: A systematic review, meta-analysis, and economic assessment. PloS One. 2021;16:e0248258.

10. Taskinen HK, Kyyrönen P, Sallmén M, et al. Reduced fertility among female wood workers exposed to formaldehyde. Am J Ind Med. 1999;36:206–212.

11. Weisskopf MG, Morozova N, O’Reilly EJ, et al. Prospective study of chemical exposures and amyotrophic lateral sclerosis. J Neurol Neurosurg Psychiatry. 2009;80:558–561.

12. Roberts AL, Johnson NJ, Cudkowicz ME, Eum K-D, Weisskopf MG. Job-related formaldehyde exposure and ALS mortality in the USA. J Neurol Neurosurg Psychiatry. 2016;87:786–788.

13. Seals RM, Kioumourtzoglou M-A, Gredal O, Hansen J, Weisskopf MG. Occupational Formaldehyde and ALS. Eur J Epidemiol. 2017;32:893–899.

14. Letellier N, Gutierrez L-A, Pilorget C, et al. Association Between Occupational Exposure to Formaldehyde and Cognitive Impairment. Neurology. 2022;98:e633–e640.

15. Protano C, Buomprisco G, Cammalleri V, et al. The Carcinogenic Effects of Formaldehyde Occupational Exposure: A Systematic Review. Cancers. 2021;14:165.

16. National Academies of Sciences, Engineering, and Medicine; Division on Earth and Life Studies; Board on Environmental Studies and Toxicology; Committee on Review of EPA’s 2022 Draft Formaldehyde Assessment. Review of EPA’s 2022 Draft Formaldehyde Assessment. Washington (DC): National Academies Press (US). 2023

17. Vaughan TL, Stewart PA, Teschke K, et al. Occupational exposure to formaldehyde and wood dust and nasopharyngeal carcinoma. Occup Environ Med. 2000;57:376–384.

18. Luce D, Leclerc A, Bégin D, et al. Sinonasal cancer and occupational exposures: a pooled analysis of 12 case-control studies. Cancer Causes Control CCC. 2002;13:147–157.

19. Schwilk E, Zhang L, Smith MT, Smith AH, Steinmaus C. Formaldehyde and leukemia: an updated meta-analysis and evaluation of bias. J Occup Environ Med. 2010;52:878–886.

20. Laforest L, Luce D, Goldberg P, et al. Laryngeal and hypopharyngeal cancers and occupational exposure to formaldehyde and various dusts: a case-control study in France. Occup Environ Med. 2000;57:767–773.

21. Beane Freeman LE, Blair A, Lubin JH, et al. Mortality from lymphohematopoietic malignancies among workers in formaldehyde industries: the National Cancer Institute Cohort. J Natl Cancer Inst. 2009;101:751–761.

22. Directorate-General for Employment, Social Affairs and Inclusion (European Commission), Klein CL, Nielsen GD, Johanson G, Bolt HM, Papameletiou D. SCOEL/REC/125 formaldehyde: recommendation from the Scientific Committee on Occupational Exposure Limits. Publications Office of the European Union. 2016.

23. Directive - 2019/983 - EN - EUR-Lex Available from: https://eur-lex.europa.eu/eli/dir/2019/983/oj/eng.

24. EH40/2005 workplace exposure limits: containing the list of workplace exposure limits for use with the Control of Substances Hazardous to Health Regulations 2002 (as amended). Fourth edition. London: TSO; 2020.

25. The GB mandatory classification and labelling list (GB MCL List) - HSE Available from: https://www.hse.gov.uk/chemical-classification/classification/mcl-list.htm.

26. The Control of Substances Hazardous to Health Regulations 2002, Section 7(7)(c)(ii) Available from: https://www.legislation.gov.uk/uksi/2002/2677/regulation/7.

27. Health and Safety Executive HSE. L5 Control of Substances Hazardous to Health: The Control of Substances Hazardous to Health Regulations 2002. Approved Code of Practice and Guidance, L5. Norwich: The Stationery Office Ltd; 2013.

28. US EPA NCFEA. IRIS Toxicological Review of Formaldehyde (Inhalation) (Final Report, 2024). US EPA Available from: https://iris.epa.gov/document/&deid%3D361799. 2024.

29. Satta G, Edmonstone J. Consolidation of pathology services in England: have savings been achieved? BMC Health Serv Res. 2018;18:862.

30. Cammalleri V, Pocino RN, Marotta D, et al. Occupational scenarios and exposure assessment to formaldehyde: A systematic review. Indoor Air. 2022;32:e12949.

31. Kangarlou MB, Dehdashti A, Saleh E. A detailed methodology for estimating health-related hazards of workplace exposure to indoor formaldehyde vapours. MethodsX. 2024;13:102937.

32. Costa S, Coelho P, Costa C, et al. Genotoxic damage in pathology anatomy laboratory workers exposed to formaldehyde. Toxicology. 2008;252:40–8.

33. Costa S, Pina C, Coelho P, et al. Occupational exposure to formaldehyde: genotoxic risk evaluation by comet assay and micronucleus test using human peripheral lymphocytes. J Toxicol Environ Health A. 2011;74:1040–51.

34. Ladeira C, Viegas S, Carolino E, Prista J, Gomes MC, Brito M. Genotoxicity biomarkers in occupational exposure to formaldehyde—The case of histopathology laboratories. Mutat Res Toxicol Environ Mutagen. 2011;721:15–20.

35. Santovito A, Schilirò T, Castellano S, et al. Combined analysis of chromosomal aberrations and glutathione S-transferase M1 and T1 polymorphisms in pathologists occupationally exposed to formaldehyde. Arch Toxicol. 2011;85:1295–1302.

36. Bouraoui S, Mougou S, Brahem A, et al. A Combination of Micronucleus Assay and Fluorescence In Situ Hybridization Analysis to Evaluate the Genotoxicity of Formaldehyde. Arch Environ Contam Toxicol. 2013;64:337–344.

37. Costa S, Carvalho S, Costa C, et al. Increased levels of chromosomal aberrations and DNA damage in a group of workers exposed to formaldehyde. Mutagenesis. 2015;30:463–73.

38. Freedom of Information Act 2000 Available from: https://www.legislation.gov.uk/ukpga/2000/36/contents.

39. Smith RL. Use of Monte Carlo simulation for human exposure assessment at a superfund site. Risk Anal Off Publ Soc Risk Anal. 1994;14:433–439.

40. BS EN 689:2018 | 30 Apr 2019 | BSI Knowledge Available from: https://knowledge.bsigroup.com/products/workplace-exposure-measurement-of-exposure-by-inhalation-to-chemical-agents-strategy-for-testing-compliance-with-occupational-exposure-limit-values.

41. Wilkins RJ, Macleod HD. Formaldehyde induced DNA—Protein crosslinks in *Escherichia coli*. Mutat Res Mol Mech Mutagen. 1976;36:11–16.

42. Ye X, Ji Z, Wei C, et al. Inhaled formaldehyde induces DNA–protein crosslinks and oxidative stress in bone marrow and other distant organs of exposed mice. Environ Mol Mutagen. 2013;54:705–718.

43. Jiménez-Villarreal J, Betancourt-Martínez ND, Carranza-Rosales P, et al. Formaldehyde induces DNA strand breaks on spermatozoa and lymphocytes of Wistar rats. Cytol Genet. 2017;51:65–73.

44. Casanova M, Morgan K, Steinhagen W, Everitt J, Popp J, Heck H. Covalent binding of inhaled formaldehyde to DNA in the respiratory tract of rhesus monkeys: pharmacokinetics, rat-to-monkey interspecies scaling, and extrapolation to man. Fundam Appl Toxicol Off J Soc Toxicol. 1991;17:409–28.

45. Crosby RM, Richardson KK, Craft TR, Benforado KB, Liber HL, Skopek TR. Molecular analysis of formaldehyde-induced mutations in human lymphoblasts and e. coli. Environ Mutagen. 1988;12:155–166.

46. Liu Y, Li C, Lu Z, Ding S, Yang X, Mo J. Studies on formation and repair of formaldehyde-damaged DNA by detection of DNA-protein crosslinks and DNA breaks. Front Biosci J Virtual Libr. 2006;11:991–7.

47. Kawanishi M, Matsuda T, Yagi T. Genotoxicity of formaldehyde: molecular basis of DNA damage and mutation. Front Environ Sci.;2:1–8.

48. Grafstrom RC, Fornace Jr, Autrup H, Lechner JF, Harris CC. Formaldehyde damage to DNA and inhibition of DNA repair in human bronchial cells. Science. 1983;220:216–8.

49. Nadalutti CA, Stefanick DF, Zhao M-L, et al. Mitochondrial dysfunction and DNA damage accompany enhanced levels of formaldehyde in cultured primary human fibroblasts. Sci Rep. 2020;10:1–13.

50. Merk O, Speit G. Significance of formaldehyde-induced DNA-protein crosslinks for mutagenesis. Environ Mol Mutagen. 1998;32:260–8.

51. Chen D, Fang L, Mei S, et al. Regulation of Chromatin Assembly and Cell Transformation by Formaldehyde Exposure in Human Cells. Environ Health Perspect. 2017;125:097019.

52. Lan Q, Smith MT, Tang X, et al. Chromosome-wide aneuploidy study of cultured circulating myeloid progenitor cells from workers occupationally exposed to formaldehyde. Carcinogenesis. 2015;36:160–167.

53. Zhang L, Tang X, Rothman N, et al. Occupational Exposure to Formaldehyde, Hematotoxicity, and Leukemia-Specific Chromosome Changes in Cultured Myeloid Progenitor Cells. Cancer Epidemiol Biomarkers Prev. 2010;19:80–88.

54. d’Ettorre G, Caroli A, Mazzotta M. Minimizing formaldehyde exposure in a hospital pathology laboratory. WORK. 2021;69:209–213.

55. Scheepers PTJ, Graumans MHF, Beckmann G, et al. Changes in Work Practices for Safe Use of Formaldehyde in a University-Based Anatomy Teaching and Research Facility. Int J Environ Res Public Health. 2018;15:2049.

56. Ogawa M, Kabe I, Terauchi Y, Tanaka S. A strategy for the reduction of formaldehyde concentration in a hospital pathology laboratory. J Occup Health. 2019;61:135–142.

57. Yahyaei E, Majlesi B, Naimi Joubani M, et al. Occupational Exposure and Risk Assessment of Formaldehyde in the Pathology Departments of Hospitals. Asian Pac J Cancer Prev APJCP. 2020;21:1303–1309.

58. Foroughi P, Golbabaei F, Sadeghi-Yarandi M, Yaseri M, Fooladi M, Kalantary S. Occupational exposure, carcinogenic and non-carcinogenic risk assessment of formaldehyde in the pathology labs of hospitals in Iran. Sci Rep. 2024;14:12006.

59. Perdelli F, Spagnolo AM, Cristina ML, et al. [Occupational exposure to formaldehyde in three pathology departments]. Ann Ig Med Prev E Comunita. 2006;18:481–490.

60. Royal College of Pathologists. Spending Review 2025 Available from: https://www.rcpath.org/static/8f6d10dd-de58-4ff5-bd339bf4999fdcbc/RCPath-submission-to-the-spending-review-2025.pdf. 2025.

61. Dugheri S, Massi D, Mucci N, Berti N, Cappelli G, Arcangeli G. How improvements in monitoring and safety practices lowered airborne formaldehyde concentrations at an Italian university hospital: a summary of 20 years of experience. Arch Ind Hyg Toxicol. 2020;71:178–189.

62. The Control of Substances Hazardous to Health Regulations 2002, Section 7(5)(a) Available from: https://www.legislation.gov.uk/uksi/2002/2677/regulation/7.

63. US EPA. Chemistry, Fate, and Transport Assessment for Formaldehyde (Final Report, 2024). US EPA. Available from: https://www.epa.gov/system/files/documents/2025-01/2.-formaldehyde-.-chemistry-fate-and-transport-assessment-.-public-release-.-hero-.-dec-2024.pdf. 2024.

64. Joaquim Pintado Nunes, Shengli Niu, Grace Monica Halim, Dafne Papandrea, Andreas Hoibl. Safe and Healthy Working Environments For All. International Labour Organization (2023).

